# Interethnic Validation of an ECG Image Analysis Software for Detecting Left Ventricular Dysfunction in Emergency Department Population

**DOI:** 10.1101/2024.10.15.24315559

**Authors:** Haemin Lee, Woon Yong Kwon, Kyoung Jun Song, You Hwan Jo, Joonghee Kim, Youngjin Cho, Ji Eun Hwang, Yeongho Choi

## Abstract

**Background:** We previously developed and validated an AI-based ECG analysis tool (ECG Buddy) in a Korean population. This study aims to validate its performance in a U.S. population, specifically assessing its LV Dysfunction Score and LVEF-ECG feature for predicting LVEF <40%, using NT-ProBNP as a comparator.

**Methods:** We identified emergency department (ED) visits from the MIMIC-IV dataset with information on LVEF <40% or ≥40%, along with matched 12-lead ECG data recorded within 48 hours of the ED visit. The performance of ECG Buddy’s LV Dysfunction Score and LVEF-ECG feature was compared with NT-ProBNP using Receiver Operating Characteristic - Area Under the Curve (ROC-AUC) analysis.

**Results:** A total of 22,599 ED visits were analyzed. The LV Dysfunction Score had an AUC of 0.905 (95% CI: 0.899 - 0.910), with a sensitivity of 85.4% and specificity of 80.8%. The LVEF-ECG feature had an AUC of 0.908 (95% CI: 0.902 - 0.913), sensitivity 83.5%, and specificity 83.0%. NT-ProBNP had an AUC of 0.740 (95% CI: 0.727 - 0.752), with a sensitivity of 74.8% and specificity of 62.0%. The ECG-based predictors demonstrated superior diagnostic performance compared to NT-ProBNP (all p<0.001).

In the Sinus Rhythm subgroup, the LV Dysfunction Score achieved an AUC of 0.913, and LVEF-ECG had an AUC of 0.917, both outperforming NT-ProBNP (0.748, 95% CI: 0.732 - 0.763, all p<0.001).

**Conclusion:** ECG Buddy demonstrated superior accuracy compared to NT-ProBNP in predicting LV systolic dysfunction, validating its utility in a U.S. ED population.

## Introduction

Accurate assessment of left ventricular (LV) dysfunction is critical in the emergency department (ED), where timely and effective treatment decisions often depend on understanding cardiac function. In conditions such as respiratory distress or shock, identifying LV dysfunction early can guide appropriate interventions.

Echocardiography is the standard for evaluating LV systolic function. However, it requires skilled operators and devices. In addition, performing it on every patients that can be benefitted from LV function information during high patient volumes in the ED can be challenging.

In recent years, artificial intelligence (AI) has become a valuable tool for analyzing electrocardiograms (ECGs) to detect conditions like myocardial infarction, heart failure, and electrolyte imbalances [1–3]. As ECGs are more readily available in the ED, AI-based ECG analysis offers a scalable alternative solution when echocardiography is not feasible.

ECG Buddy, an AI-driven ECG analysis tool, can assess various emergencies and cardiac function abnormalities using 12-lead ECG images. Previous studies have demonstrated that ECG Buddy outperforms clinical experts in diagnosing critical conditions such as ST-elevation myocardial infarction (STEMI), hyperkalemia, and right ventricular (RV) dysfunction [4–7]. Additionally, its evaluation of left ventricular (LV) systolic function has been shown to outperform NT-ProBNP and match the accuracy of point-of-care ultrasound (POCUS) performed by emergency physicians [8].

However, its prior external validation studies have primarily been conducted in a Korean population [4–11], leaving a gap in evidence regarding its performance in populations with different racial profiles, particularly non-East Asian groups. This limitation raises the need to investigate its utility in broader populations.

This study aims to validate ECG Buddy’s performance in a U.S. population, particularly its ability to predict Heart Failure with reduced Ejection Fraction (HFrEF) as defined by left ventricular ejection fraction (LVEF) <40%, and to compare its diagnostic accuracy with NT-ProBNP, a standard biomarker used in heart failure assessment.

## Materials and methods

### Study Design

This study is a retrospective analysis designed to validate the performance of ECG Buddy, an AI-based ECG analysis tool, in predicting reduced LV systolic dysfunction (LVEF <40%) in a U.S. population. Data was obtained from the MIMIC-IV (Medical Information Mart for Intensive Care-IV) dataset (v2.2) and its associated ECG dataset (MIMIC-IV-ECG v1.0), which contains comprehensive de-identified health records and ECG waveform dataset from patients admitted to the emergency departments (EDs) and critical care units of the Beth Israel Deaconess Medical Center [12–13].

### Study Population and Data Collection

The process of selecting ED visit events for this study involved multiple stages of filtering from the MIMIC-IV dataset (Fig. 1). Briefly, we started by filtering 331,784 discharge notes using a regular expression search for echocardiography-related terms, reducing it to 97,929 records. Next, the most recent GPT-4o-mini model (24-08-20) was used to label each note based on three key questions: whether an echocardiogram was performed during the admission, whether the patient underwent heart-related surgeries or procedures, and whether there was evidence of LVEF <40% (Supplementary Table S1). The dataset was then reduced to 33,780 records by selecting those for patients who had an echocardiogram performed during the admission and did not undergo heart-related surgeries or
procedures, based on the answers to the first two questions provided by the language model. We then selected a single ECG for each visit that are most chronologically near to the time of hospital arrival from the ECGs performed between 24 hours before and 72 hours after the hospital arrival for each patient, further reducing the dataset to 26,042 cases. An emergency medicine specialist then reviewed the answer to the third question provided by the model to correct or confirm the presence of LV dysfunction (LVEF <40%). During this process, cases with unknown LV dysfunction status were excluded, leaving 24,947 cases. Finally, only ED visits were included, resulting in a final dataset of 22,599 ED patient visits.

**Fig 1.**
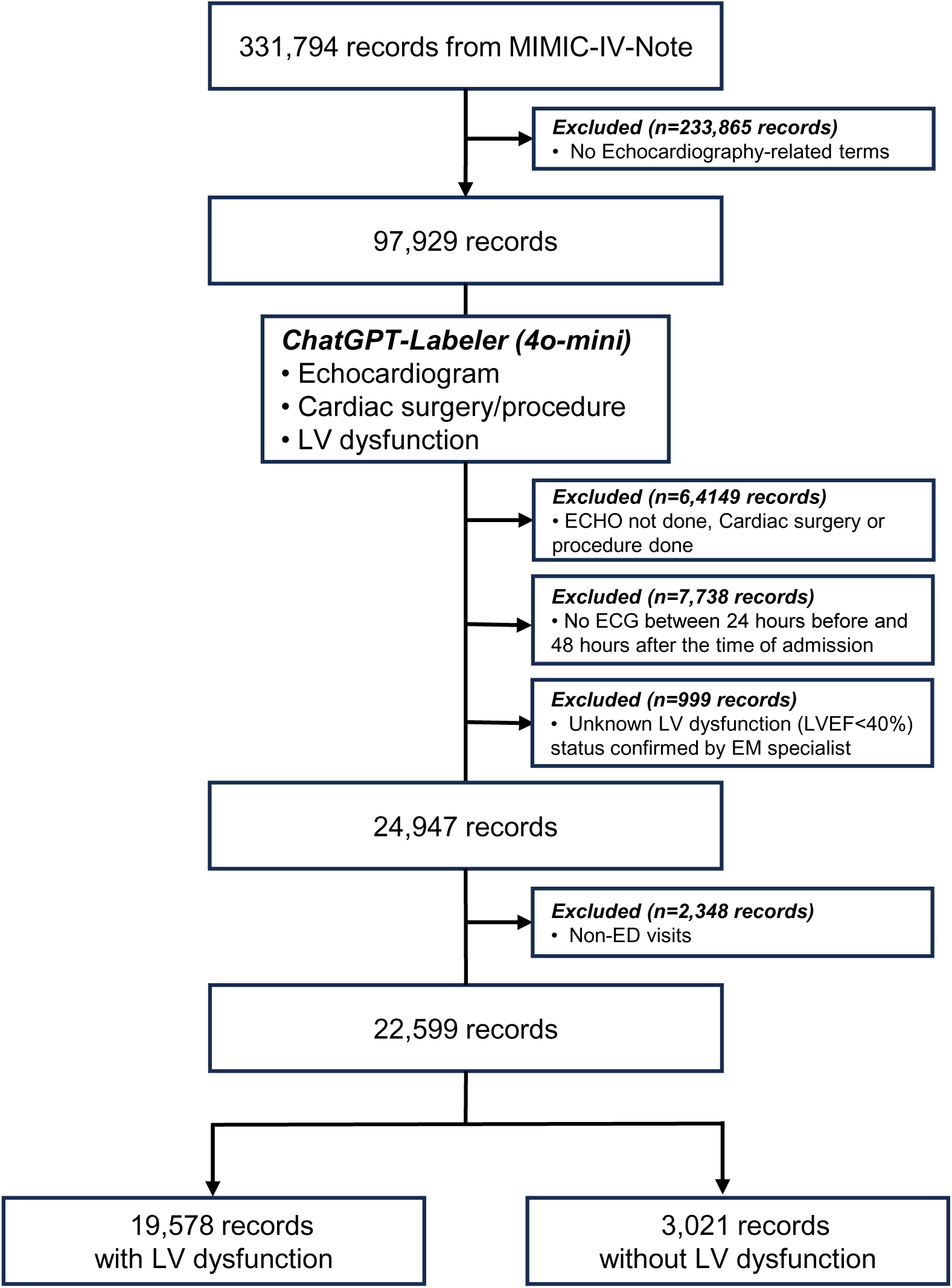
Data Selection Process from MIMIC-IV Dataset.; MIMIC-IV: Medical Information Mart for Intensive Care-IV; ECG: electrocardiogram; LV: left ventricular; EM: emergency medicine; ED: emergency department

### Extraction of ECG Biomarkers using ECG Buddy

ECG Buddy is a deep learning-based AI platform that analyzes 12-lead ECG images, available via smartphones, desktop PCs, or direct EHR integration. It classifies heart rhythms and generates 10 digital biomarkers, termed Quantitative ECG (QCG) scores, for a range of emergencies, cardiac dysfunctions, and hyperkalemia [4–7]. It has been approved as a Class II medical device in South Korea and is available in various app stores. The developers have recently added a new biomarker, LVEF-ECG, which estimates the patient’s LVEF directly. This feature is not yet available for public use, as it is currently under evaluation for MFDS approval in Korea. In this study, we used the research version of ECG Buddy for Windows, which allows batch analysis of large ECG datasets for both of the biomarkers.

We extracted LV Dysfunction score (a risk score for LV systolic dysfunction, which is LVEF<40), ranging 0 to 100 and LVEF-ECG ranging from 20 to 65 to test their performance in identifying reduced LVEF (LVEF <40%) as determined from the review of discharge notes.

### Statistical Analysis

The performance of the software for screening LV dysfunction was assessed using area under the Receiver Operating Characteristic curve (AUC-ROC). Initial NT-ProBNP level within 72 hours for each of the ED visit were used as comparator. Sensitivity and specificity were determined by binning the predictors using the thresholds maximizing their Youden’s index. A subgroup analysis was conducted for patients with sinus rhythms (normal sinus rhythm, sinus bradycardia and sinus tachycardia) to assess the software’s performance without being influenced by arrhythmia. Statistical significance was established at a P-value of <0.05, and R software version 4.1.0 was used for data analysis [14].

### Ethical Considerations

The study used de-identified data from the MIMIC-IV database, which is publicly available for research purposes and complies with HIPAA standards for de-identification. No patient consent was required due to the retrospective nature of the study and the use of anonymized data.

## Results

A total of 22,599 ED visits were analyzed. Patients were divided into two groups based on their LV dysfunction status. Baseline characteristics of the two groups are presented in Table 1.

**Table 1.** Patient Characteristics.

|  | **No LV Dysfunction**<br>**LVEF≥40 (N = 19,578)** | **LV Dysfunction**<br>**LVEF<40 (N = 3,021)** | **p-value** |
| --- | --- | --- | --- |
| **Age (years)** | 72.0 (59.0, 83.0) | 75.0 (63.0, 84.0) | <0.001 |
| **Gender** |  |  | <0.001 |
| - Female | 10,566 (54.0%) | 1,071 (35.5%) |  |
| - Male | 9,012 (46.0%) | 1,950 (64.5%) |  |
| **Race** |  |  | <0.001 |
| - White | 13,343 (68.2%) | 1,997 (66.1%) |  |
| - Black/African American | 3,566 (18.2%) | 668 (22.1%) |  |
| - Hispanic/Latino | 1,001 (5.1%) | 149 (4.9%) |  |
| - Asian | 585 (3.0%) | 57 (1.9%) |  |
| - Native/Other | 59 (0.3%) | 5 (0.2%) |  |
| - Unknown | 1,024 (5.2%) | 145 (4.8%) |  |
| **Chest pain** |  |  | 0.048 |
| - No | 11,104 (87.4%) | 1,739 (85.7%) |  |
| - Yes | 1,606 (12.6%) | 289 (14.3%) |  |
| **Dyspnea** |  |  | <0.001 |
| - No | 9,469 (74.5%) | 1,347 (66.4%) |  |
| - Yes | 3,241 (25.5%) | 681 (33.6%) |  |
| **Fever** |  |  | 0.009 |
| - No | 12,152 (95.6%) | 1,965 (96.9%) |  |
| - Yes | 558 (4.4%) | 63 (3.1%) |  |
| **Temperature (°C)** | 36.7 (36.4, 37.0) | 36.6 (36.3, 36.9) | <0.001 |
| **Heart rate (bpm)** | 83.0 (70.0, 98.0) | 86.0 (72.0, 100.0) | 0.001 |
| **Respiratory rate (bpm)** | 18.0 (16.0, 20.0) | 18.0 (16.0, 20.0) | <0.001 |
| **O2 saturation (%)** | 98.0 (96.0, 100.0) | 98.0 (96.0, 100.0) | 0.823 |
| **Systolic BP (mmHg)** | 136.0 (118.0, 155.0) | 127.0 (110.0, 145.0) | <0.001 |
| **Diastolic BP (mmHg)** | 74.0 (63.0, 86.0) | 74.0 (63.0, 86.0) | 0.693 |
| **Time to ECG (min)** | 1,450.0 (1,217.0, 1,717.0) | 1,421.0 (1,194.0, 1,686.0) | <0.001 |
| **1-week mortality** |  |  | <0.001 |
| - No | 19,263 (98.4%) | 2,908 (96.3%) |  |
| - Yes | 315 (1.6%) | 113 (3.7%) |  |
| **1-month mortality** |  |  | <0.001 |
| - No | 18,524 (94.6%) | 2,703 (89.5%) |  |
| - Yes | 1,054 (5.4%) | 318 (10.5%) |  |
| **LV Dysfunction Score** | 0.101 (0.033, 0.274) | 0.752 (0.502, 0.887) | <0.001 |
| **LVEF-ECG (%)** | 57.6 (51.3, 60.8) | 33.6 (27.3, 42.9) | <0.001 |

Patients with LV dysfunction were older on average (median age of 75.0 years vs. 72.0; p<0.001) and more likely to be male (64.5% vs. 46.0%, p<0.001). The racial composition was predominantly White in both groups, though the proportion of Black/African American patients was slightly higher in the LVEF <40% group (22.1% vs. 18.2%, p<0.001). Dyspnea was more common in the LV dysfunction group (33.6% vs. 25.5%, p<0.001).

Fig. 2 and Table 2 shows the performance of the ECG biomarkers. In the overall population, the LV Dysfunction Score generated by ECG Buddy had an AUC of 0.905 (95% CI: 0.899 - 0.910). The sensitivity was 85.4% and specificity was 80.8%, with a threshold of 0.352 determined by the Youden index. The LVEF-ECG feature had an AUC of 0.908 (95% CI: 0.902 - 0.913), with a sensitivity of 83.5% and specificity of 83.0% using a threshold of 47.178. In comparison, NT-ProBNP had an AUC of 0.740 (95% CI: 0.727 - 0.752), with a sensitivity of 74.8% and specificity of 62.0%, at a threshold of 2828.5.

**Fig 2.**
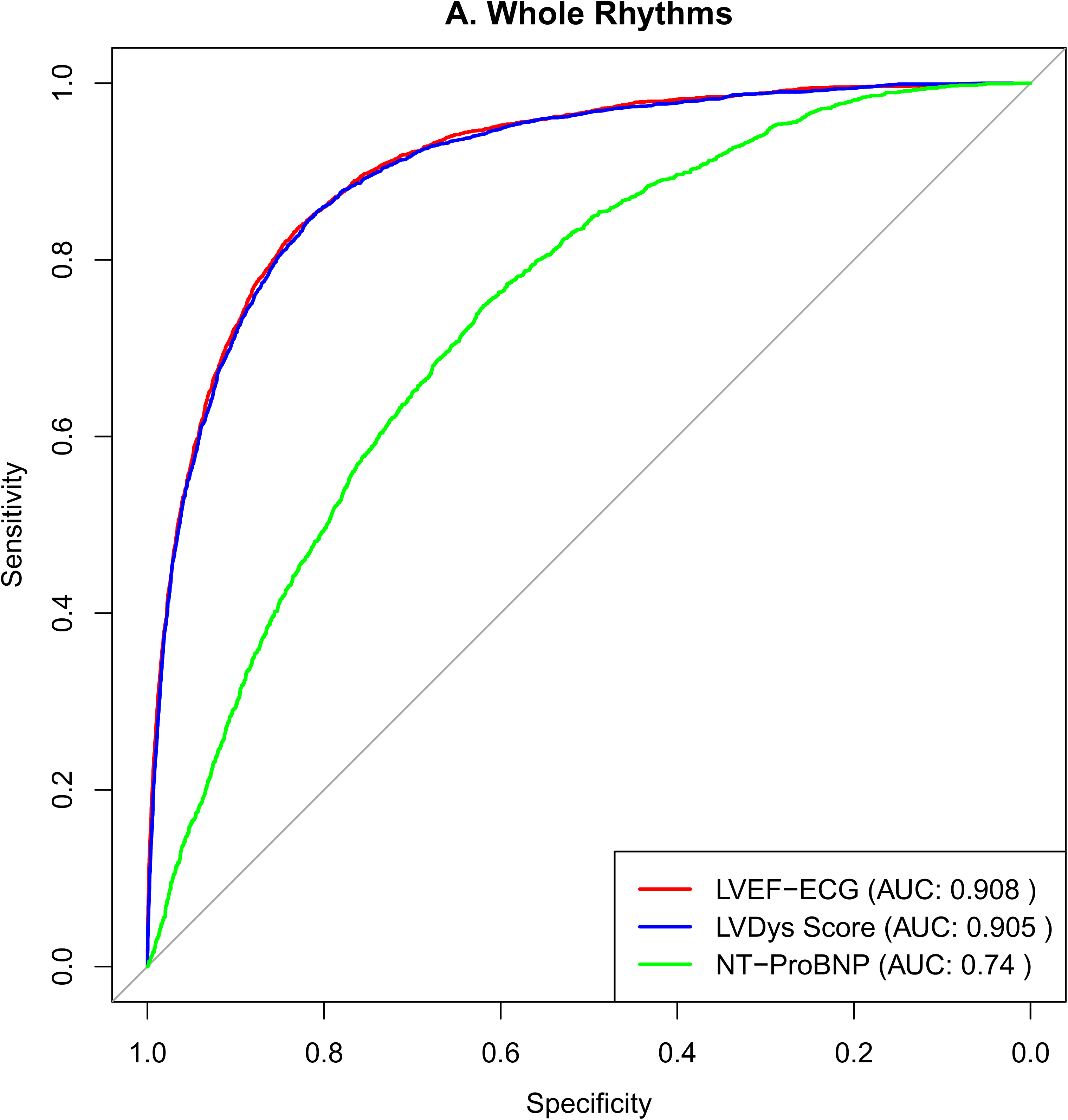
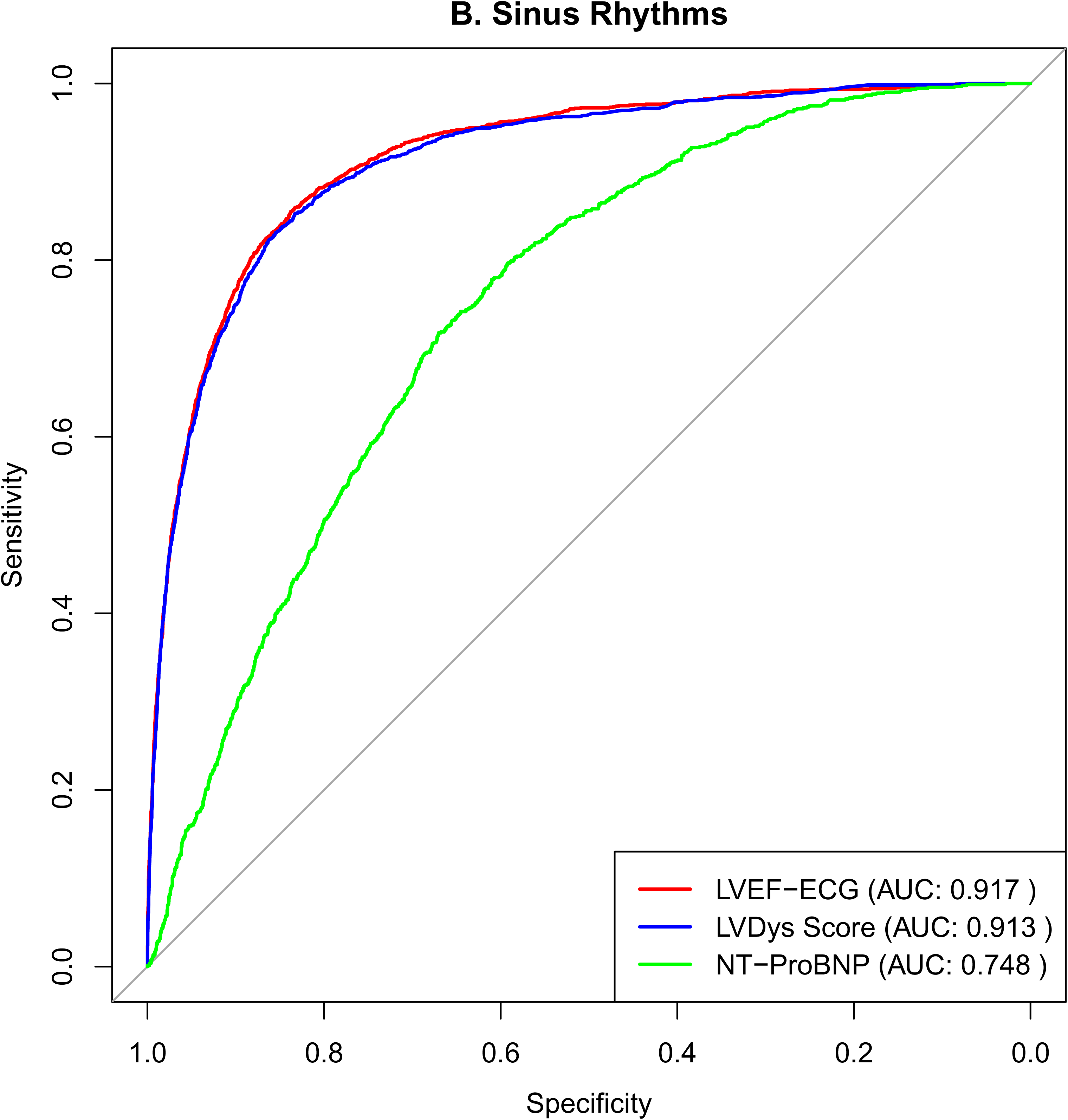
Receiver Operating Characteristic Curves for the digital biomarkers of ECG Buddy.; A: Whole rhythm population, B: Sinus rhythm population

**Table 2.** Performance Summary.

| Population | Predictor | AUC (95% CI) | Threshold | Sensitivity | Specificity |
| --- | --- | --- | --- | --- | --- |
| All Rhythms | LV Dysfunction Score | 0.905 (0.899 - 0.910) | 0.352 | 85.4% | 80.8% |
|  | LVEF-ECG | 0.908 (0.902 - 0.913) | 47.178 | 83.5% | 83.0% |
|  | NT-ProBNP | 0.740 (0.727 - 0.752) | 2828.5 | 74.8% | 62.0% |
| Sinus Rhythm | LV Dysfunction Score | 0.913 (0.906 - 0.920) | 0.352 | 83.1% | 85.5% |
|  | LVEF-ECG | 0.917 (0.910 - 0.924) | 49.138 | 85.5% | 83.8% |
|  | NT-ProBNP | 0.748 (0.732 - 0.763) | 1796.5 | 79.7% | 59.2% |

In the Sinus Rhythm subgroup (N=16,527), the LV Dysfunction Score achieved an AUC of 0.913 (95% CI: 0.906 - 0.920), with a sensitivity of 83.1% and specificity of 85.5%, with a threshold of 0.352. The LVEF-ECG feature had an AUC of 0.917 (95% CI: 0.910 - 0.924), with a sensitivity of 85.5% and specificity of 83.8%, at a threshold of 49.138. NT-ProBNP’s performance in the Sinus Rhythm subgroup showed an AUC of 0.748 (95% CI: 0.732 - 0.763), with a sensitivity of 79.7% and specificity of 59.2%, using a threshold of 1796.5.

In both the overall population and the Sinus Rhythm subgroup, ECG Buddy demonstrated superior diagnostic performance compared to NT-ProBNP (all p<0.001).

## Discussion

In this study, we evaluated the diagnostic accuracy of two ECG-based AI biomarkers, LV Dysfunction Score and LVEF-ECG, in comparison to NT-ProBNP for detecting left ventricular systolic dysfunction in ED patients. Both ECG-based biomarkers outperformed NT-ProBNP in identifying patients with LVEF <40%. This marks the first demonstration of ECG Buddy’s accuracy in a U.S. population, highlighting the potential of its digital biomarkers to serve as effective diagnostic tools for LV dysfunction in emergency settings, offering a more accessible and rapid alternative to traditional methods like NT-ProBNP.

Previous studies have shown ECG Buddy’s ability to outperform clinical experts in screening conditions such as STEMI, hyperkalemia, and RV dysfunction using 12-lead ECGs. Additionally, its performance to screen LV dysfunction has been shown to be on par with point-of-care ultrasound (POCUS) performed by emergency physicians [4–9]. However, these studies were primarily conducted in a Korean population. One of the strengths of this study is the inclusion of a U.S. population with a broader racial profile, making it more relevant to diverse clinical settings. This validation in a more diverse population provides new evidence of ECG Buddy’s generalizability across different racial and ethnic groups.

In the fast-paced environment of ED, timely and accurate identification of LV dysfunction is critical for guiding treatment decisions. While echocardiography is the gold standard for assessing LVEF, it can be challenging to perform on all patients due to logistical constraints, especially during periods of high patient volume. As a result, NT-ProBNP is often used to screen for heart failure in EDs. However, NT-ProBNP is neither particularly accurate nor cost-effective. In contrast, ECG Buddy’s AI-driven approach offers a more accurate and scalable solution, particularly in emergency settings where ECGs are already required to evaluate multiple differential diagnoses, such as arrhythmias and myocardial ischemia. Additionally, its image-based analysis enables easy and rapid deployment through smartphones or PCs, making implementation in real-world settings straightforward.

In this study, we leveraged a language model (GPT-4o-mini) to perform large-scale analysis of discharge notes. With over 97,929 discharge notes to review, manual inspection would have been extremely time-consuming and resource-intensive. By utilizing GPT-4o-mini, we efficiently filtered the dataset down to approximately 26,000 cases, significantly reducing the workload and enabling a more manageable review process. While some errors were observed in determining LVEF <40%, necessitating manual chart reviews, this combination of automated filtering and selective manual review proved highly effective in handling large clinical dataset.

One key limitation of this study is the reliance on discharge notes to determine LVEF. The timing of echocardiography relative to the ECG in the emergency department is unclear, potentially introducing variability, especially since cardiac function can change during hospitalization due to disease progression and treatments. While we used GPT-4o to exclude cases with documented surgeries or procedures, some variability remains.

## Conclusions

This study demonstrates that ECG Buddy’s AI-driven digital biomarkers, LV Dysfunction Score and LVEF-ECG, provide superior diagnostic accuracy compared to NT-ProBNP for identifying LV systolic dysfunction in a U.S. ED population. As the first validation of ECG Buddy in a racially diverse U.S. population, the findings confirm its utility as a reliable tool for detecting LV dysfunction across different clinical and demographic groups. Further studies are encouraged to validate these results in other healthcare settings and explore its broader clinical applications.

## Data Availability

All data used in this study are publicly available and can be accessed from MIMIC-IV, available online at https://physionet.org/content/mimiciv/2.2/.

## Acknowledgments

This research was supported by a grant of the Korea Health Technology R&D Project through the Korea Health Industry Development Institute (KHIDI), funded by the Ministry of Health & Welfare, Republic of Korea (grant number : RS-2023-00265933), and the SNUBH Research Fund (grant number: 14-2023-0335).

## Author contributions

1. Conceptualization: Haemin Lee, Ji Eun Hwang, Yeongho Choi
2. Data curation: Haemin Lee, Ji Eun Hwang, Yeongho Choi
3. Formal analysis: Haemin Lee, Ji Eun Hwang, Yeongho Choi
4. Funding acquisition: Joonghee Kim, Youngjin Cho, Ji Eun Hwang, Yeongho Choi, Woon Yong Kwon, Kyoung Jun Song, You Hwan Jo
5. Investigation: Haemin Lee, Ji Eun Hwang, Yeongho Choi
6. Methodology: Haemin Lee, Ji Eun Hwang, Yeongho Choi, Woon Yong Kwon, Kyoung Jun Song, You Hwan Jo
7. Project administration: Haemin Lee
8. Resources: Haemin Lee, Joonghee Kim, Youngjin Cho
9. Software: R software version 4.1.0
10. Supervision: Ji Eun Hwang, Yeongho Choi, Woon Yong Kwon, Kyoung Jun Song, You Hwan Jo
11. Validation: Ji Eun Hwang, Yeongho Choi
12. Visualization: Haemin Lee
13. Writing – Haemin Lee
14. Writing – review & editing: Ji Eun Hwang, Yeongho Choi, Woon Yong Kwon, Kyoung Jun Song, You Hwan Jo
15. Approval of final manuscript: all authors

## References

[1] Cho Y, Kwon J, Kim KH, Medina-Inojosa J, et al. Artificial intelligence algorithm for detecting myocardial infarction using six-lead electrocardiography. Scientific reports, 2020, 10.1: 20495.

[2] Zhao Y, Xiong J, Hou Y, et al. Early detection of ST-segment elevated myocardial infarction by artificial intelligence with 12-lead electrocardiogram. International Journal of Cardiology, 2020, 317: 223–230.

[3] Makimoto H, Höckmann M, Lin T, et al. Performance of a convolutional neural network derived from an ECG database in recognizing myocardial infarction. Scientific reports, 2020, 10.1: 8445.

[4] Kim D, Hwang JE, Cho Y, et al. A retrospective clinical evaluation of an artificial intelligence screening method for early detection of STEMI in the emergency department. Journal of Korean medical science, 2022, 37.10.

[5] Kim D, Jeong J, Kim J, et al. Hyperkalemia Detection in Emergency Departments Using Initial ECGs: A Smartphone AI ECG Analyzer vs. Board-Certified Physicians. Journal of Korean Medical Science, 2023, 38.45.

[6] Choi YJ, Park MJ, Cho Y, et al. Screening for RV Dysfunction Using Smartphone ECG Analysis App: Validation Study with Acute Pulmonary Embolism Patients. Journal of Clinical Medicine, 2024, 13.16: 4792.

[7] Choi YJ, Park MJ, Ko Y, et al. Artificial intelligence versus physicians on interpretation of printed ECG images: Diagnostic performance of ST-elevation myocardial infarction on electrocardiography. International journal of cardiology, 2022, 363: 6–10.

[8] Kim JH, Kim J, Cho Y, et al. Non-Inferiority Analysis of Mobile ECG Analyzer vs. POCUS for Screening Left Ventricular Dysfunction in the Emergency Department. 대한응급의학회 학술대회 초록집. 2023, 2023.2: 1–3.

[9] Choi HM, Cho Y, Kim J, et al. ECG-derived global longitudinal strain using artificial intelligence: A comparative study with transthoracic echocardiography. Journal of the American College of Cardiology, 2024, 83.13_Supplement: 2360–2360.

[10] Cho Y, Yoon M, Kim J, et al. Artificial Intelligence–Based Electrocardiographic Biomarker for Outcome Prediction in Patients With Acute Heart Failure: Prospective Cohort Study. Journal of medical Internet research, 2024, 26: e52139.

[11] Park MJ, Choi YJ, Shim M, et al. Performance of ECG-Derived Digital Biomarker for Screening Coronary Occlusion in Resuscitated Out-of-Hospital Cardiac Arrest Patients: A Comparative Study between Artificial Intelligence and a Group of Experts. Journal of Clinical Medicine, 2024, 13.5: 1354.

[12] Johnson A, Bulgarelli L, Pollard T, et al. MIMIC-IV (version 2.2). PhysioNet. 2024. 10.13026/hxp0-hg59.

[13] Johnson A, Bulgarelli L, Shen L, et al. MIMIC-IV, a freely accessible electronic health record dataset. Sci Data, 2023, 10.1. 10.1038/s41597-022-01899-x

[14] R: A language and environment for statistical computing. R Foundation for Statistical Computing, Vienna, Austria. https://www.R-project.org/.

