## Supplemental Table 1 for "Interethnic Validation of an ECG Image Analysis Software for Detecting Left Ventricular Dysfunction in Emergency Department Population"

**Supplementary**

**Table S1. GPT-4o-mini (24-08-20) Prompt design**

'Instructions:

Analyze the discharge note and respond to the following questions. Provide your answers in JSON format. Use "yes", "no", or "unknown" as the value for each key. Regardless of whether LV dysfunction is confirmed or not, quote the part of the discharge note describing the ejection fraction (EF) or relevant heart function information.

Answer "yes" for LV dysfunction confirmation only if:

The ejection fraction is explicitly stated as below 40%, or

A range is given with the upper margin below 40%, or

the (global not regional) LV dysfunction is described as severe. (excluding moderate to severe severity)

Answer "no" if:

The lower margin of the ejection fraction range is 40% or higher, or

The echocardiogram or LV function is described as "normal," "within normal limits," or indicates "no problems.", or

the (global not regional) LV dysfunction is described as mild.(excluding mild to moderate severity)

Answer "unknown" if:

No exact number is provided, and the description is ambiguous (e.g., "reduced EF" without specific numbers), or

The ejection fraction falls within an uncertain range (e.g., 30-40%), or

described as "moderate", "mild to moderate" or "moderate to severe".

Always quote the part of the note that describes the ejection fraction or heart function.

Questions:

Was an echocardiogram conducted during this hospitalization?

Was any cardiac surgery or procedure performed during this hospitalization?

Did the echocardiogram confirm LV dysfunction (Left ventricular ejection fraction below 40%)? Follow the rules above for ranges, qualitative descriptions, and unknown cases.

Output format:

{"echocardiogram_performed": "yes/no/unknown",

"cardiac_surgery_or_procedure": "yes/no/unknown",

"lv_dysfunction_confirmed": {

"answer": "yes/no/unknown",

"description": "quoted text from discharge note"} }'
